# Gut microbial communities modulate efficacy of albendazole-ivermectin against soil-transmitted helminthiases

**DOI:** 10.1101/2021.10.07.21264625

**Authors:** Pierre H.H. Schneeberger, Morgan Gueuning, Sophie Welsche, Eveline Hürlimann, Julian Dommann, Cécile Häberli, Jürg E. Frey, Somphou Sayasone, Jennifer Keiser

## Abstract

**Background:** Soil-transmitted helminth infections represent a large burden across the globe with over a quarter of the world’s population at risk. The outcome of available treatments is species-specific with a large proportion of unexplained treatment failure. Administration of albendazole is the standard of care, but because of low cure rates (CR) observed in treating *Trichuris trichiura* infections, a significantly more efficacious alternative therapy combining albendazole and ivermectin is being investigated.

**Methods:** 80 patients from the village of Pak-Khan, in Laos, with confirmed STH infections (*Trichuris trichiura* and hookworms), received either albendazole (400 mg) or albendazole (400 mg) and ivermectin (200 µg/kg) together. A pre-treatment stool sample was collected as well as daily post-treatment stool samples for up to 28 days to measure treatment efficacy. Taxonomic profiling of pre-treatment stool samples was conducted using 16S rRNA gene sequencing, target-specific and total bacteria qPCR, as well as shotgun sequencing.

**Results:** Three bacterial communities, or enterotypes (ET) 1-3, were identified. No association with pre-treatment enterotype and treatment outcome of both *Trichuris trichiura* and hookworm were found in the monotherapy arm with overall cure rates (CR) of 7.5% and 50%, respectively. Pre-treatment enterotype was strongly associated with efficacy of the combination therapy for both, *T. trichiura* (CR_overall_ = 33.3%; CR_ET1_ = 5.8%; CR_ET2_ = 16.6%; CR_ET3_ = 68.5%) and hookworm (CR_overall_ = 47.2%; CR_ET1_ = 31.2%; CR_ET2_ = 16.6%; CR_ET3_ = 78.5%) infections. Daily post-treatment egg per gram of stool counts recapitulated these observations and faster and increased egg reduction was observed in ET3 when compared to failure-associated ET1 and ET2. Species-level comparisons of these enterotypes highlighted a set of ten differentially enriched bacterial species.

**Conclusion:** Taxonomically distinct gut microbiota communities were found in this setting in terms of both, relative and absolute abundances, of specific bacterial taxa. Pre-treatment enterotype was relevant for treatment outcome of the combination therapy, albendazole and ivermectin, for *T. trichiura* as well as for hookworm infections. These observations indicate that pre-treatment microbial composition of stool samples should be monitored to ensure evidence-based administration of albendazole-ivermectin to treat these diseases.

## Introduction

Over 25% of the world’s population is at risk being of infected by soil-transmitted helminth (STH)^1^. *Ascaris lumbricoides, Trichuris trichiura* and hookworms (*Ancylostoma duodenale* and *Necator americanus*) account for most STH infections and the current recommended treatment is mainly based on two benzimidazole drugs, albendazole and mebendazole, which display varying species-specific treatment efficacies^2^. For instance, a single-dose regimen of either drug will achieve poor cure rates treating *T. trichiura* infections^3,4^ while displaying high efficacy against *A. lumbricoides* infections^5^. Moreover, mass drug administration (MDA) campaigns, which represent the main control strategy^6^, can lead to increased selective pressure towards resistant parasites. Yet, the drug pipeline for new drug candidates remains empty despite recent efforts to discover novel drugs^7^, and relies mainly on repurposing existing drugs from veterinary medicine^8^. The use of drug combinations, e.g. albendazole with ivermectin, has recently been advertised to broaden the spectrum of activity of treatments against soil-transmitted helminthiasis^9^. Ivermectin is a derivate of avermectin, which was initially isolated in *Streptomyces avermectinius*^10^. It possesses a 16-membered macrocyclic lactone group and thus belongs to the macrolide drug class, which also encompasses antibiotic drugs such as azithromycin and erythromycin, used to treat mostly gram-positive bacterial infections. Ivermectin is commonly used in the veterinary field and has been shown to improve efficacy of single-dose treatment against *T. trichiura* when administered in combination with albendazole^11^. However, in a recent multi-country randomized controlled trial, varying treatment efficacy of albendazole-ivermectin against *T. trichiura* was observed with cure rates ranging from 8% to 66% in different settings^12^.

One potential confounder of drug treatment efficacy is the dense and diverse non-parasitic gut microbiome^13^. Its pivotal role in treatment outcome is currently being investigated in different fields, including cancer research and metabolic diseases^14,15^. However, there is still very limited information pertaining to its role in the context of neglected tropical diseases and more specifically how it affects treatment efficacy of essential drugs such as benzimidazoles for STH infections^16,17^. Various mechanisms of drug-microbe interactions have been identified but can broadly be categorized into either direct or indirect mechanisms^13^. Direct metabolization of drugs by gut microbes most often result in modulation of potency and/or toxicity is a widespread mechanism^18^. Indirect mechanisms include, but are not limited to, local microbe-driven mechanisms of tampered gut wall function, resulting in deregulated drug translocation into systemic circulation^19,20^.

In this study, we aimed to identify potential inter-kingdom mechanisms of resilience (i.e. = bacteria modulating anti-parasitic drug efficacy) in the treatment of *T. trichiura* and hookworm infections. Understanding possible interactions between gut microbes and anthelminthic drugs could help to improve and optimize current treatment efficacy while avoiding selection towards resistant microbial communities. In the framework of this randomized trial, we assessed efficacy of two anthelminthic treatments, albendazole alone (400 mg), and in combination with ivermectin (200 µg/kg), in the context of gut microbial communities. We conducted taxonomic profiling of gut microbial communities on pre-treatment stool samples to predict treatment efficacy assessed by post-treatment egg reduction rates. Microbial community composition was determined using several techniques, including 16S rRNA gene sequencing, 16S- and taxon-specific qPCR, as well as shotgun sequencing to identify species-level taxonomic features associated with treatment outcome.

## Results

### Sampling results and study characteristics

Between January and May 2019, people from ten villages in Nambak, Luang Prabang district in Laos were tested for *T. trichiura*. 549 people were included and randomly assigned to either monotherapy (albendazole) or combination therapy (albendazole-ivermectin)^21^. Daily post-treatment stool samples were collected from a subset of 88 patients up to 28 days after first administration, among which 80 were included for microbiome assessment (**Figure 1**). Among these, 86.2% (n=69) were also co-infected with hookworms, 40% (n=32) with *A. lumbricoides*, and 8.7% (N=7) with *Opisthorchis viverrini*. Treatment arms were balanced in terms of sex, age, baseline infection intensities for any helminths, and no noteworthy between-group differences were observed (**Table 1**). Similarly, sampling compliance in the post-treatment period was comparable between both treatment arms (**Figure S1**).

**Figure 1.**
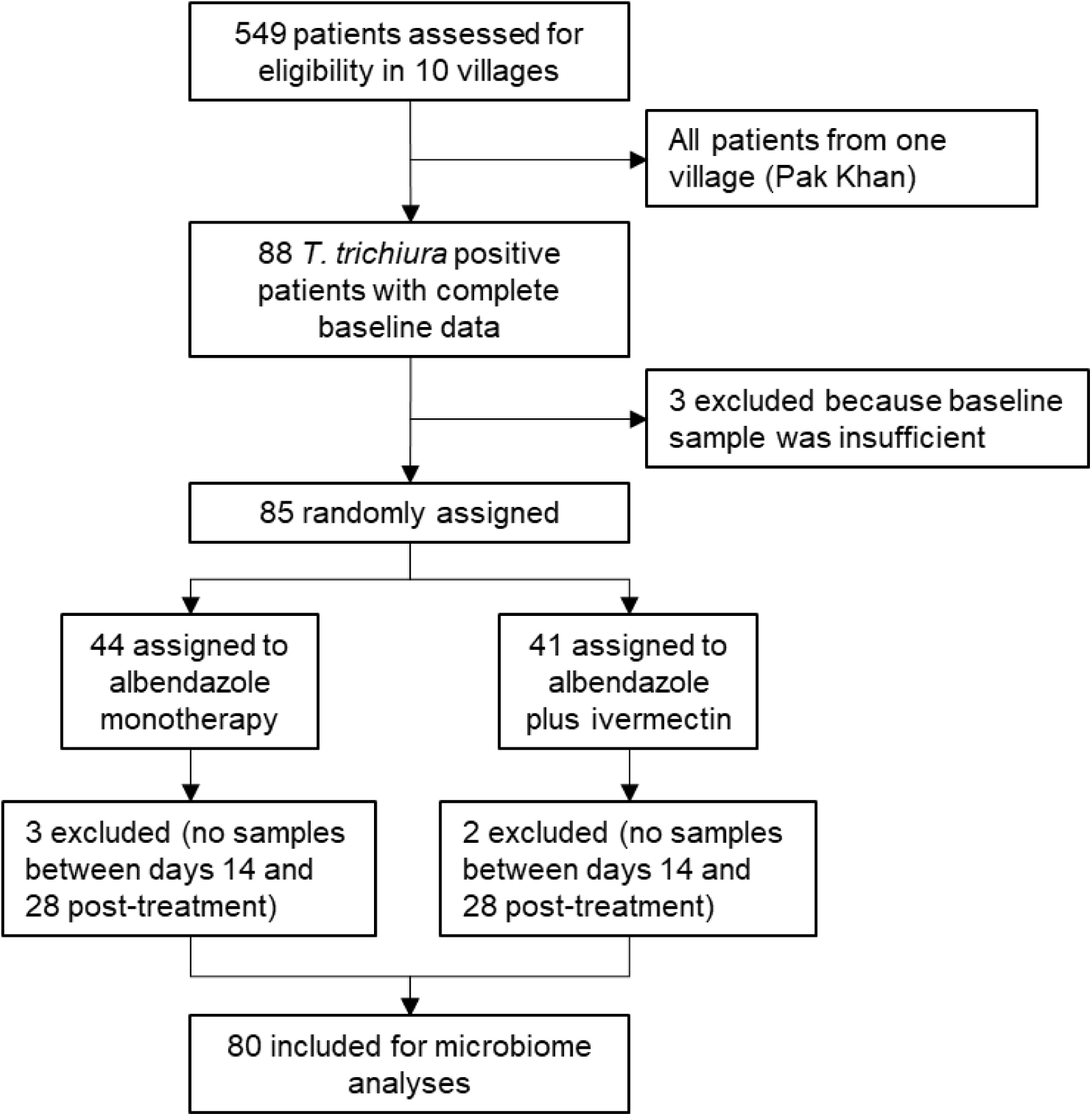
Trial profile.

**Table 1.** Cohort description. y = year; ATX = antibiotic treatment within one month before treatment; EPG = eggs per gram of stool; SD = standard deviation; arit = arithmetic mean; geom = geometric mean; I.i = Infection intensity based on World Health Organization criteria.

| Characteristic | Albendazole-ivermectin | Albendazole | P-value |
| --- | --- | --- | --- |
| Number of patients (n) | 39 | 41 |  |
| Age (y) |  |  |  |
| Mean (arit) (± SD) | 29 (± 17) | 26 (± 17) | 0.37 |
| Range | 6-56 | 6-60 |  |
| Female, n (%) | 25 (64.1%) | 20 (48.8%) | 0.18 |
| **ATX (< 1 month)** | 0 (0%) | 2 (4.8%) | 0.49 |
| ***Trichuris trichiura* infection** |  |  |  |
| Infected, n (%) | 39 (100%) | 41 (100%) |  |
| EPG<sub>arit</sub> (± SD) | 1001 (± 1427) | 793 (± 884) | 0.45 |
| EPG<sub>geom</sub> (± SD) | 497 (± 3.1) | 501 (± 2.6) |  |
| Range (EPG) | 102-6054 | 114-5004 |  |
| I.i: Light, n (%) | 28 (71.8%) | 30 (73.1%) |  |
| I.i: Moderate, n (%) | 11 (28.2%) | 11 (26.8%) |  |
| **Hookworm infection** |  |  |  |
| Infected, n (%) | 35 (89.7%) | 34 (82.9%) |  |
| EPG<sub>arit</sub> (± SD) | 1418 (± 1777) | 1548 (± 3861) | 0.86 |
| EPG<sub>geom</sub> (± SD) | 766 (± 3.2) | 358 (± 5.9) |  |
| Range (EPG) | 54-8574 | 12-22416 |  |
| I.i: Light, n (%) | 28 (80%) | 27 (79.4%) |  |
| I.i: Moderate, n (%) | 4 (11.4%) | 4 (11.8%) |  |
| I.i: Heavy, n (%) | 3 (8.6%) | 3 (8.8%) |  |
| ***Ascaris lumbricoides* infection** |  |  |  |
| Infected, n (%) | 15 (38.5%) | 17 (41.5%) |  |
| EPG<sub>arit</sub> (± SD) | 7992 (± 8481) | 6739 (± 9961) | 0.72 |
| EPG<sub>geom</sub> (± SD) | 2558 (± 9.3) | 1423 (± 10.2) |  |
| Range (EPG) | 6-26256 | 12-38922 |  |
| I.i: Light, n (%) | 8 (50%) | 12 (70.5%) |  |
| I.i: Moderate, n (%) | 8 (50%) | 5 (29.5%) |  |
| ***Opisthorchis viverrini* infection** |  |  |  |
| Infected, n (%) | 2 (5.1%) | 5 (12.2%) |  |
| EPG<sub>arit</sub> (± SD) | 15 (± 3) | 39 (± 35) | 0.45 |
| EPG<sub>geom</sub> (± SD) | 766 (± 3.2) | 358 (± 5.9) |  |
| Range (EPG) | 12-18 | 6-108 |  |

### Compositional clustering and cluster-specific taxonomic features

Unsupervised clustering based on taxonomic composition before treatment enabled identification of underlying community structures into three distinct clusters, or enterotypes (ET1-3), as shown in **Figure 2A**. The proportion represented by each enterotype was comparable between both treatment arms (**Figure S2**). Accuracy of this enterotype-based classification was 78.75% with a Kappa statistic of 62.93% when assessed using a random forest model. Most important taxonomic features in terms of classification accuracy included the genera *Faecalibacterium* (2.704%), *Escherichia/Shigella* complex (2.613%), *Prevotella* (1.814%), *Phascolarctobacterium* (1.082%), and [*Eubacterium*] *coprostanoligenes* group (1.044%) (**Figure 2B**). Relative abundances of *Faecalibacterium* and *Prevotella* were significantly higher in ET1 (*P* = 2.61E-09 and 7.55E-05, respectively), *Escherichia/Shigella* in ET2 (*P* = 1.33E-07), and [*Eubacterium*] *coprostanoligenes* group was enriched in ET3 (*P* = 7.55E-05) among a set of eight genera found to be differentially abundant between the three enterotypes (**Figure 2C**). Taxonomic composition was homogeneous when comparing baseline samples from both treatment arms with PERMANOVA (r^2^ = 0.013, *P* = 0.38), as shown in **Figure 2D**.

**Figure 2.**
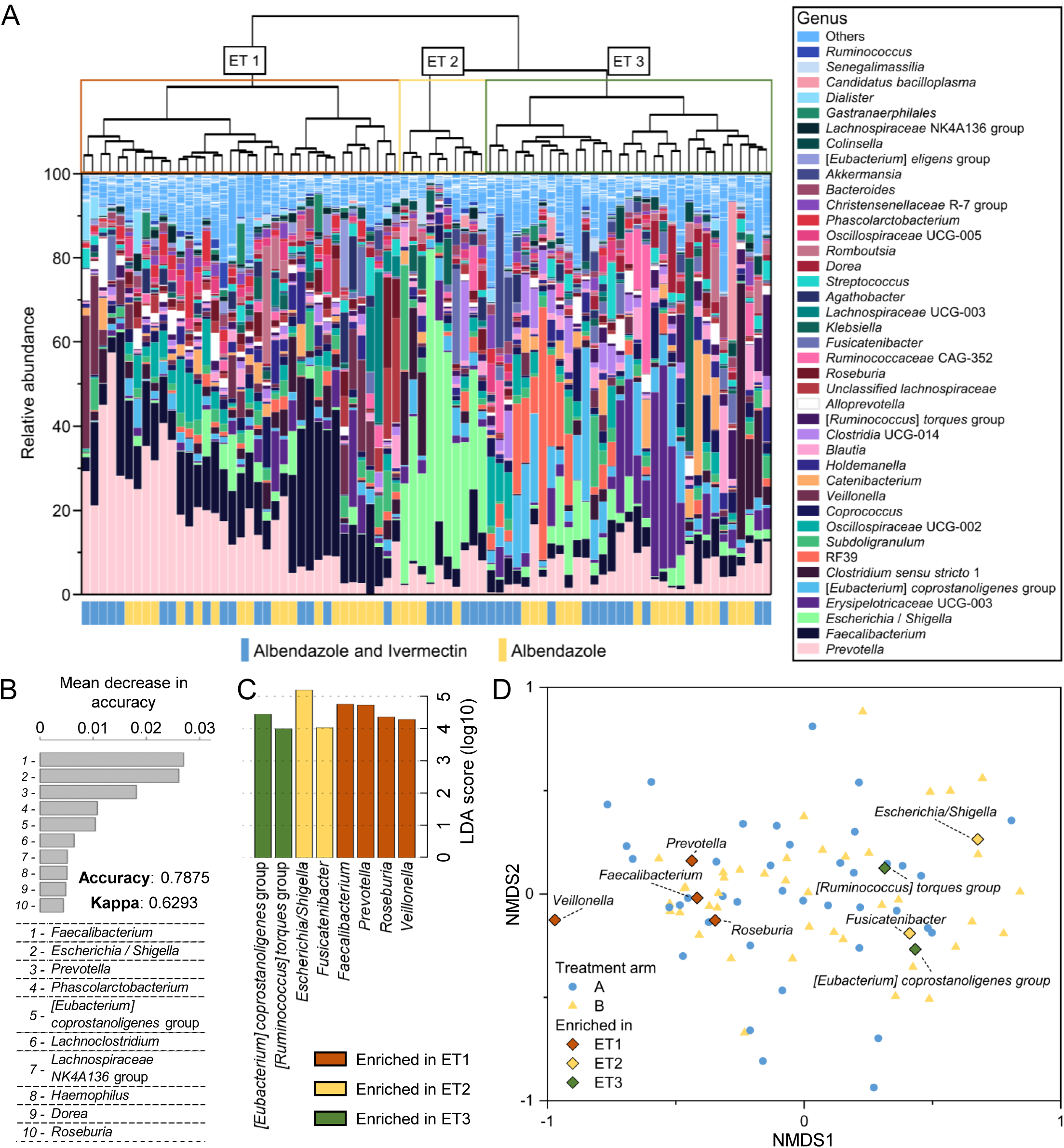
Underlying compositional structures and their taxonomic features. **A**. Gut microbial community composition of patients infected with Trichuris trichiura before treatment. The cladogram was generated using Bray-Curtis dissimilarity. **B**. Performance of classification in this dataset using a random forest model. Taxonomic features are ranked according to their individual contribution to sample classification. **C**. Genera found to be enriched in one of the enterotype using a Kruskal-Wallis test for group comparison combined with a linear discriminant analysis for effect size (Lefse). **D**. Non-metric multidimensional scaling (NMDS) ordination plot of baseline samples using Bray-Curtis distance. The labelled genera were found to be enriched in either enterotype. ALB+IVE = albendazole and ivermectin; ALB = albendazole; ET = Enterotype.

### Taxon-specific absolute abundances and total bacteria confirm presence of different community structures

Absolute abundances of *Faecalibacterium* (FAEC), *Escherichia* (ESCH), and *Prevotella* (PREV) assessed through qPCR confirmed the 16S rRNA gene sequencing-based enterotype classification (**Figure 3A**). For instance, ESCH was significantly higher in ET2 than in both ET1 and ET3 (*P* = 0.001 and 0.002). Similarly, FAEC and PREV were both significantly higher (*P* = 0.001 and *P* = 0.004, respectively) in ET1 than in ET3. We then tested whether qPCR values obtained from these three targets and total bacterial load (16S) could be used to discriminate between these microbial signatures. The qPCR-based random forest model accurately classified 83.1% of samples (Kappa = 0.72) into enterotypes 1-3, which is similar to the metrics obtained with the sequencing-based classification. Area under the receiver operating curve (AUC) of this model were of 1 (95%CI = N.A) between ET1 and ET2, of 0.88 (95%CI = 0.79-0.97) between ET1 and ET3, and of 0.91 (95%CI = 0.82-1) between ET2 and ET3 (**Figure 3B**) indicating the suitability of qPCR-based classification into an identified enterotype as an alternative to high-throughput sequencing.

**Figure 3.**
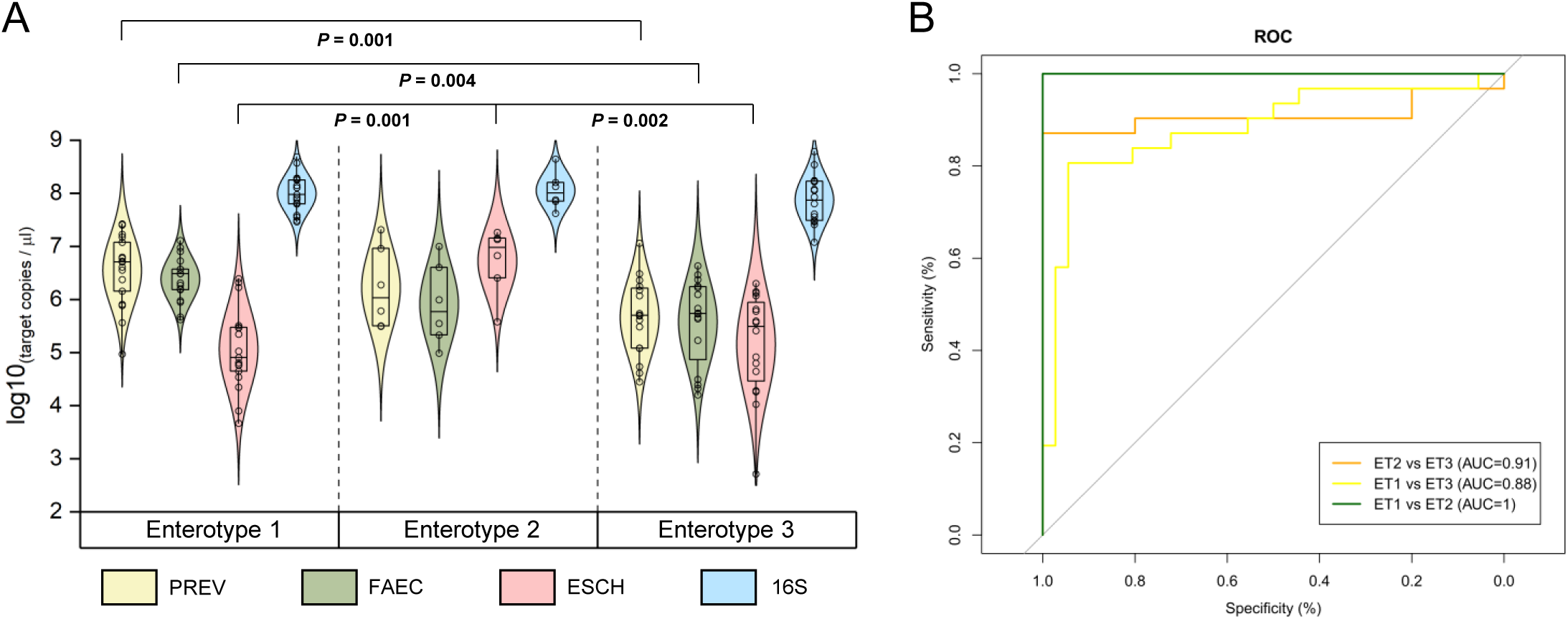
Taxon-specific and total bacterial qPCR to classify pre-treatment sample in a treatment-relevant category. **A**. Total bacteria and taxon-specific density measured by quantitative PCR (qPCR), by enterotype. **B**. Classification sensitivity and specificity into the enterotype using qPCR values. PREV = *Prevotella* genus qPCR target; FAEC = *Faecalibacterium* genus qPCR target; ESCH = Escherichia coli qPCR target; 16S = total bacteria qPCR target.

### Association of identified enterotype with T. trichiura and hookworm cure

We did not observe any association between cure rate, defined as an average egg per gram count (EPG) equal to zero between days 14-28 post-treatment, and ET, for patients receiving albendazole monotherapy for *T. trichiura* (**Figure 3A**, right half) nor hookworm infections (**Figure 3B**, right half). However, the majority of patients who received the combination therapy (albendazole-ivermectin) and were cured for any soil-transmitted helminth infection, presented a gut microbial composition classified as ET3 pre-treatment. This observation was similar for both, *T. trichiura* (**Figure 3A**, left half) and hookworm infections (**Figure 3B**, left half). Likelihood of cure of *T. trichiura* infection was significantly higher (*P* = 0.0002) for patients presenting ET3 than those presenting ET1 at baseline with an odds ratio (OR) of 0.03 (95%CI = 0.01-0.2). A similar, albeit weaker association (*P* = 0.025), with an OR of 0.14 (95% CI = 0.03-0.66), was observed between ET and treatment outcome of hookworm. Absolute abundances of PREV and FAEC measured at baseline correlated with average egg counts between days 14-28 post-treatment for the combination therapy with spearman correlation coefficients of 0.416 (*P* = 0.009) and 0.275 (*P* = 0.09), respectively (**Supplementary figure 3A**). No correlation was observed between *T. trichiura* cure and absolute microbial abundances in the monotherapy arm, nor for hookworm in both treatment arms (**Supplementary figure 3B-D**).

### Species-level features enriched in either failure- or success-associated enterotype in the albendazole-ivermectin treatment arm

To identify species-level differences between failure-associated enterotypes 1-2 and success-associated enterotype 3, we generated shotgun sequencing data from samples collected within the albendazole plus ivermectin treatment arm. In addition to previously observed enriched features from *Prevotella* (*P. copri* in ET1), *Faecalibacterium* (*F. prausnitzii* in ET1), *Escherichia/Shigella* (*E. coli*, in ET2), and *Ruminococcus* (*R. torques*, in ET3), we found *Streptococcus salivarius, Coprococcus eutactus, Roseburia faecis, Dorea longicatena*, and *Eubacterium halii* to be significantly enriched in either ET1 or ET3 when comparing both groups (**Figure 3C**). *Coprococcus eutactus* was also depleted in ET3 when comparing it to ET2 while *Anaerostipes hadrus* was enriched in the former.

### Cox proportional hazard models of treatment outcome stratified by pre-treatment enterotype

With two consecutive samples within the 28-days post-treatment period without eggs detected as the definition of cure of *T. trichiura* and hookworms, measured hazard ratios indicate faster egg clearance while presenting ET3 (HR = 2.89, *P* = 0.005), when compared to ET1, for patients receiving the combination therapy. This association remains true when adjusting for potential confounders such as sex, age, and infection intensities (**Table 2**). Faster clearance is also observed for hookworm cure for ET3 (HR = 2.41, *P* = 0.016) when compared to egg clearance of patients presenting ET1. This association remains true when adjusting for sex, (*P* = 0.04), infection intensity (*P* = 0.017), and age (*P* = 0.003). Probability of early egg clearance was not different between patients presenting ET2 and ET1 for both, *T. trichiura* and hookworm infections, with the combination therapy. We did not observe any association between enterotype and probability of being cured in the monotherapy arm.

**Table 2.** Prediction of time to worm clearance using pre-treatment (pre-tx) enterotypes (ET). Clearance is defined as two consecutive samples with no eggs (*Trichuris trichiura* or hookworm) detected and an average egg count between day 14 and 28 equal to zero. ALB-IVE = Albendazole and Ivermectin; ALB = Albendazole; ref. = reference category; HR (95% CI) = Hazard ratio (95% confidence interval); tx = treatment.

| *Trichuris trichiura* – ALB-IVE |  |  |  |  |  |  |
| --- | --- | --- | --- | --- | --- | --- |
| Outcome | Predictor | HR (95% CI) - ET pre-tx | P-value | Predictor | HR (95% CI) - ET pre-tx | P-value |
| Time to cure | ET 2 | 1.09 (0.18-47.06) | 0.445 | ET 3 | 2.89 (2.33-141.04) | 0.005 |
| (ref = ET1) | Adjusted for |  |  | Adjusted for |  |  |
| ALB+IVE | Sex | 1.49 (0.27-74.90) | 0.298 | Sex | 3.39 (3.48-258.04) | 0.001 |
|  | Age | 1.21 (0.19-56.35) | 0.403 | Age | 2.99 (2.46-162.89) | 0.005 |
|  | Infection intensity | 0.87 (0.15-38.43) | 0.536 | Infection intensity | 2.93 (2.39-145.98) | 0.005 |

**Table 2. Prediction of time to worm clearance using pre-treatment (pre-tx) enterotypes (ET).**
| n=39, events=13 |  |  |  |  |  |  |
| --- | --- | --- | --- | --- | --- | --- |
| *Trichuris trichiura* - ALB |  |  |  |  |  |  |
| Outcome | Predictor | HR (95% CI) - ET pre-tx | P-value | Predictor | HR (95% CI) - ET pre-tx | P-value |
| **Time to cure** | **ET 2** | 1.58 (0.16-15.20) | 0.692 | **ET 3** | 0.37 (0.04-3.57) | 0.391 |
| (ref = ET1) | Adjusted for |  |  | Adjusted for |  |  |
| **ALB** | Sex | 1.87 (0.18-19.60) | 0.601 | Sex | 0.37 (0.04-3.57) | 0.391 |
|  | Age | 1.16 (0.09-14.81) | 0.908 | Age | 0.31 (0.03-3.27) | 0.327 |
|  | Infection intensity | 2.37 (0.25-22.84) | 0.455 | Infection intensity | 0.37 (0.04-3.43) | 0.372 |
| n=41, events=5 |  |  |  |  |  |  |
| Hookworm – ALB+IVE |  |  |  |  |  |  |
| Outcome | Predictor | HR (95% CI) - ET pre-tx | P-value | Predictor | HR (95% CI) - ET pre-tx | P-value |
| **Time to cure** | **ET 2** | 0.53 (0.06-4.56) | 0.565 | **ET 3** | **2.41 (1.28-10.98)** | **0.016** |
| (ref = ET1) | Adjusted for |  |  | Adjusted for |  |  |
|  | Sex | 0.49 (0.06-4.43) | 0.529 | Sex | **3.45 (1.06-11.28)** | **0.04** |
|  | Age | 1.05 (0.11-10.2) | 0.969 | Age | **5.67 (1.76-18.28)** | **0.003** |
|  | Infection intensity | 0.56 (0.07-4.83) | 0.599 | Infection intensity | **3.71 (1.26-10.91)** | **0.017** |
| n=36, events=17 |  |  |  |  |  |  |
| Hookworm – ALB |  |  |  |  |  |  |
| Outcome | Predictor | HR (95% CI) - ET pre-tx | P-value | Predictor | HR (95% CI) - ET pre-tx | P-value |
| **Time to cure** | **ET 2** | 1.55 (0.31-7.71) | 0.592 | **ET 3** | 1.69 (0.59-4.75) | 0.322 |
| (ref = ET1) | Adjusted for |  |  | Adjusted for |  |  |
|  | Sex | 1.57 (0.31-7.81) | 0.584 | Sex | 1.88 (0.63-5.59) | 0.257 |
|  | Age | 1.62 (0.62-8.7) | 0.577 | Age | 1.78 (0.52-6.05) | 0.357 |
|  | Infection intensity | 1.55 (0.31-7.75) | 0.591 | Infection intensity | 1.67 (0.59-4.7) | 0.332 |
| n=34, events=23 |  |  |  |  |  |  |

### Associations between success- and failure-associated enterotypes and daily post-treatment eggs per gram of stool counts

In a survival analysis, we confirm that patients presenting ET3 pre-treatment are more likely to be both, faster and more efficiently cured from a *T. trichiura* infestation using the albendazole and ivermectin-based treatment than those presenting ET1 and ET2 (*P* = 0.0002; **Figure 5A**, left-half). The same observation applies in the context of hookworm infections (*P* = 0.009; **Figure 5A**, right-half). Patients with ET3 revealed the highest cure rates of 78% at 14-days post-treatment whereas patients with ET1 and ET2 reached only 31% and 16% cure in this time frame, respectively. Daily egg patterns also largely reflect these findings (**Figure 5B**). A sharp decrease of EPG counts is observed immediately following treatment, irrespective of treatment arm or baseline enterotype. For *T. trichiura* EPG counts, this decrease was more pronounced in treatment arm A (−98.2% average EPG at day 5) than arm B (−72.9% average EPG at day 5) which correlates with increased efficacy observed for the combination therapy at the recommended follow-up examination (days 14-28). Notably, EPG counts increase after the initial drop for patients with ET1, and to a lesser extent, patients with ET2, for both types of infections. There was no difference when comparing EPG count decrease for hookworms between treatment arms with an average decrease of 99.1% and 99.3% at day 5 for treatment arm A and B, respectively.

**Figure 4.**
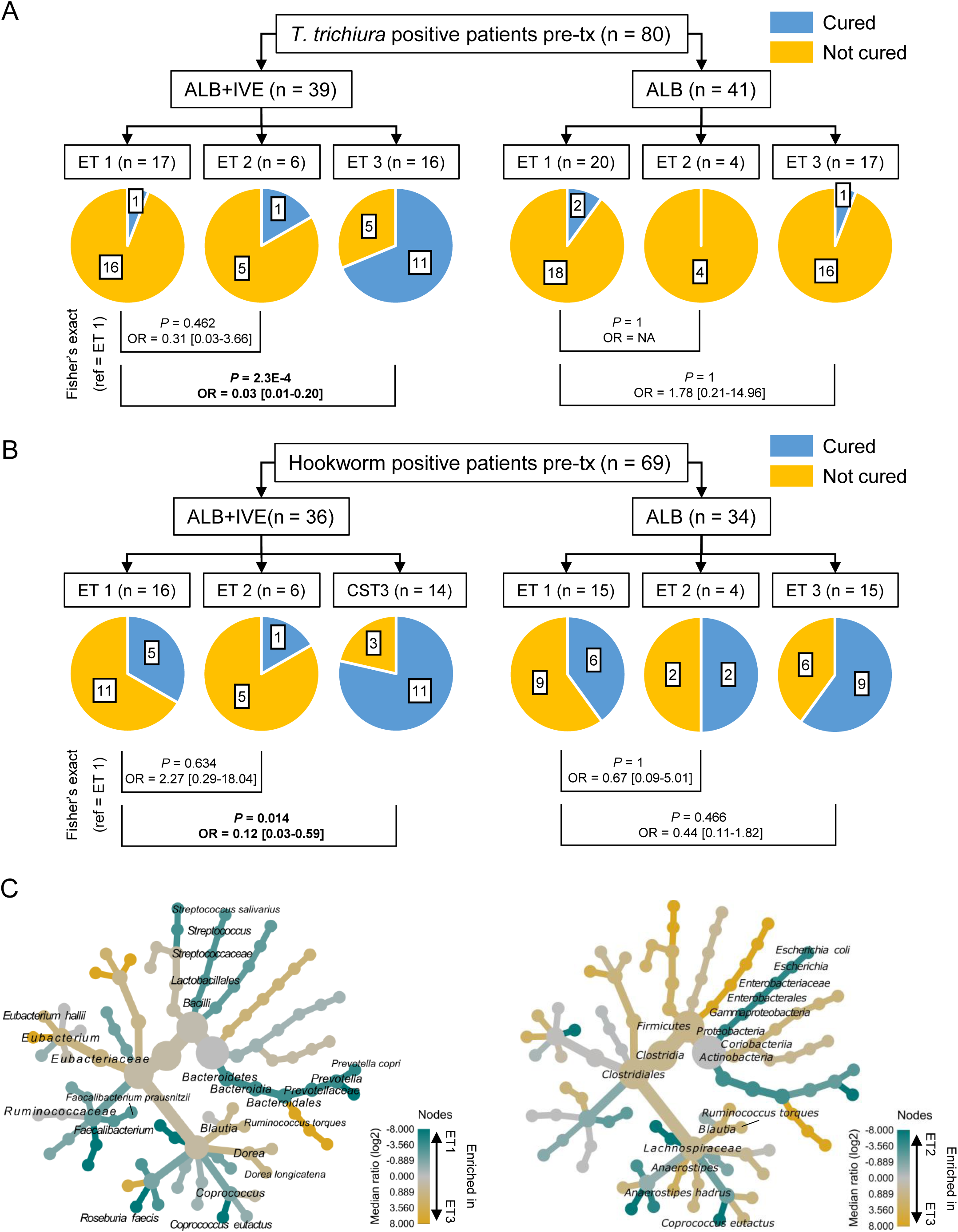
Association between soil-transmitted helminths cure and pre-treatment enterotype (ET) by treatment arm and species-level characteristics of each enterotype. **A**. Association between treatment outcome of *Trichuris trichiura* and ET at baseline. **B**. Association measured between treatment outcome of hookworm and ET before treatment. Cure rate is defined as the presence or absence of eggs in stool between day 14 and day 28 after treatment. **C**. Species-level differences between compositional clusters (= enterotypes). The 95% confidence interval of the odd ratios is shown in bracket. Labels on the pie charts represent the number of patients in each group. n = number of patients; OR = odds ratios.

**Figure 5.**
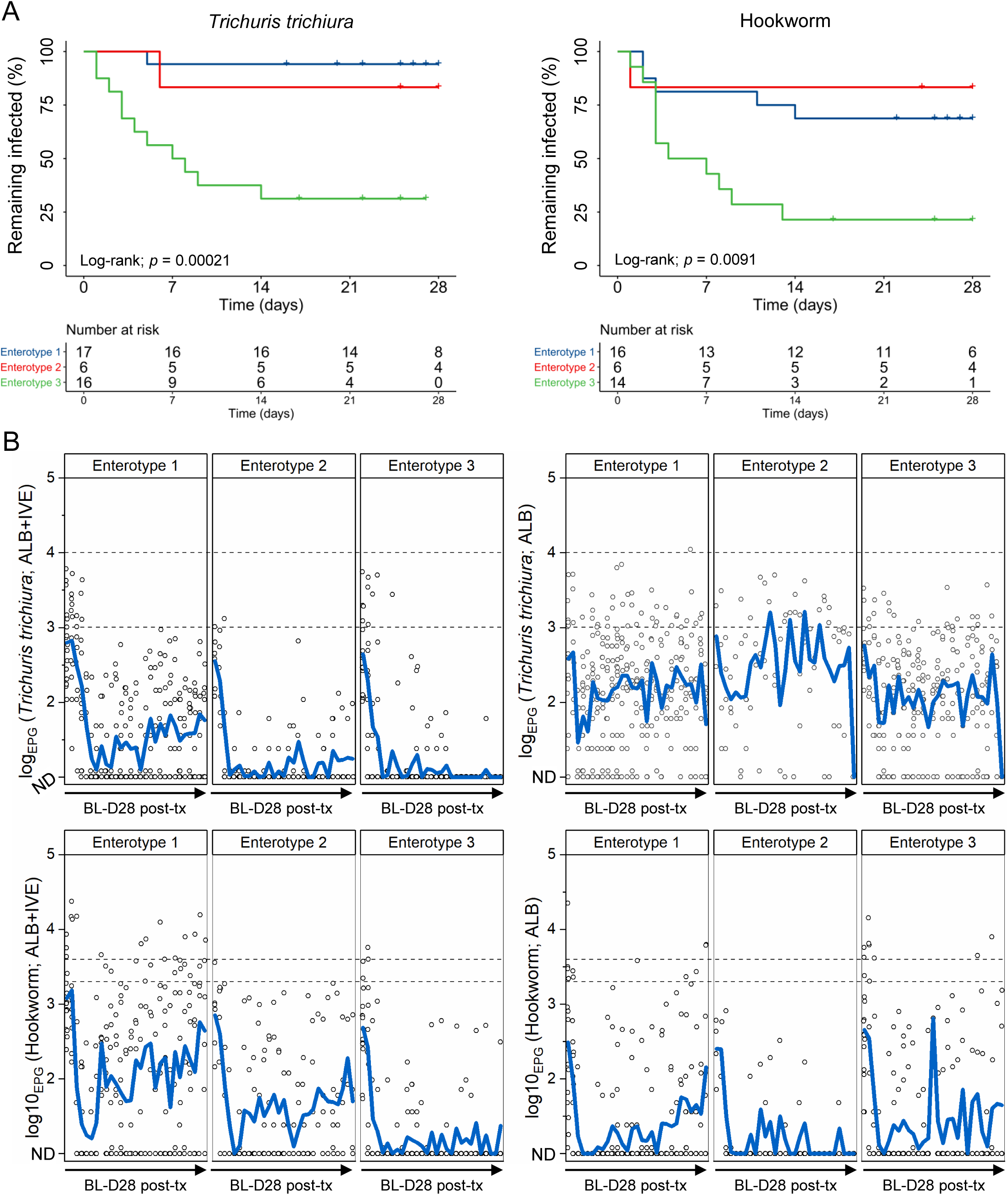
Relationship between pre-treatment enterotype and daily post-treatment egg counts of *Trichuris trichiura* and hookworm. **A**. Kaplan-Meier curve showing the remaining proportion of infected patients over time, stratified by baseline enterotype. **B**. Averaged daily egg counts of *Trichuris trichiura* and hookworm stratified by treatment arm and pre-treatment enterotype. The black dashed lines indicate the lower and upper threshold of moderate infections for each parasite. ND = not detected; post-tx = post-treatment; ALB = albendazole; IVE = ivermectin.

## Discussion

This study is the first to investigate gut microbial determinants of antiparasitic treatment failure for essential drugs used to treat human *T. trichiura* and hookworm infections. Our results represent a significant improvement of our understanding of gut microbiota-drug interaction in the context of anthelmintics treatment. The key findings are i) that taxonomically distinct communities were found in this setting in terms of both, relative and absolute abundance of specific taxa, and ii) that pre-treatment community composition, or enterotype, is associated with treatment outcome of the combination therapy, albendazole and ivermectin, for *T. trichiura* as well as for hookworm infections. These observations indicate that pre-treatment microbial composition of stool samples is strongly correlated with treatment efficiency of both *T. trichiura* and hookworms when using ivermectin-based treatment.

Ivermectin is a functionally and chemically complex drug. For instance, recent literature described the re-positioning of the drug to treat a wide range of off-label non-parasitic diseases^22,23^. As consequence, this could lead to an increase of ivermectin consumption worldwide de facto making understanding of its interactions with the gut microbiota even more relevant for public health. Furthermore, it harbours a 16-membered macrocyclic lactone group, which is structurally similar to that found in important macrolide antibiotics including, erythromycin (14-membered lactone), azithromycin (15-membered lactone), and tyrosin/josamycin (16-membered lactone), among others^24-27^. While this remains to be functionally validated, it raises the question as to whether drug resistance interactions could occur between bacteria and ivermectin, which could be driven through a potential bacteriostatic/bactericidal activity on failure-associated taxa. Further studies should investigate this direct mechanism, since several of the failure-associated taxa are gram-positive bacteria, which are known targets of macrolide antibiotics and could hence be sensitive to this drug^26^. This is, for instance, the case of *S. salivarius*. Several streptococci species have been shown to harbour multiple mechanisms of macrolide resistance, including drug efflux^28,29^, target alteration through methylation (protection) of the 23S/50S complex^29^, and perhaps even more importantly in this context, to inactivate macrolide drugs through a set of drug inactivation enzymes including macrolide phosphotransferases and erythromycin esterases^30,31^. Drug inactivation through bacterial enzymes could decrease the amount of active compound reaching their indicated targets - soil-transmitted helminths, and hence reduce overall treatment efficacy. Other potential mechanisms of interest identified in *S. salivarius* include, for instance, bioaccumulation/sequestration of drugs in bacterial cells, as recently published^32^. However, these hypotheses remain to be proven, and functional validation remains to be performed to understand mechanisms involved in this drug-microbe interaction. It is worth noting that this interaction is present for hookworms, which colonize the small intestine, as well as for *T. trichiura*, which resides in the caecum, at the beginning of the large intestine. This speaks in favour of a more widespread interaction, rather than a locally restricted event. Another key focus point of future research would also include understanding how many, among the 34-92% of non-responders to albendazole/ivermectin-based treatment, can be attributed to potential bacteria-driven interactions. This information would help define how treatment response could be improved through a microbiome-based therapy. Therapeutic opportunities derived from these findings could, for instance, involve targeted disruption of hypothetically resistant strains or overall community displacement with fitter, macrolide-sensitive strains to mitigate resistance potential. The other important implication of this study is that ivermectin, which is marketed as an antiparasitic drug, might also contribute to selecting cross-resistant bacterial isolates, which could hinder efforts involved in antibiotic stewardship programs. This is particularly important since it is widely used in MDA campaigns. Provided this resistance interaction is functionally validated, ivermectin and a related derivative moxidectin should also be considered in antimicrobial stewardship efforts. This could be achieved by using pre-treatment screening of genetic/phenotypic determinants of resistance.

However, our study has several important limitations. First, our observations are based on a cohort with relatively low sample size (n = 80). Hence, data driven beta diversity-based classification might change in a larger cohort. In addition, the taxonomic determinants of failure identified in this setting might be specific to the village of Pak-Khan in Lao PDR and might not be generalizable. While we confirmed robustness of our taxonomic-related observations with different techniques, further studies in different settings are needed to pinpoint common microbial/genetic denominators of low albendazole plus ivermectin-based treatment efficacy. Second, we did not investigate intrinsic worm resistance. While there is currently no known target for screening of *T. trichiura* and hookworm resistance to ivermectin, we cannot exclude that lower sensitivity of the parasites might explain, in part, decreased treatment efficacy. While this could be the case, our efficacy-related observations (66% failure) are in line with previous observations of failure associated with this drug combination. Since there is no evidence of parasitic resistance to ivermectin in human populations to date, we believe that the study setting is representative of an environment with normal parasitic sensitivity.

In conclusion, we showed in this study that the gut microbiota composition is an important driver of response to anthelminthic treatment based on albendazole and ivermectin. These findings will enable understanding of the cause of albendazole and ivermectin-based treatment failure in a large proportion of patients, while still being one of the most efficacious options in terms of success to treat STH infections. This will hopefully lead to novel therapeutic opportunities based on the modulation of failure-associated features, and - perhaps more importantly – to optimized, evidence-based use of these powerful drugs.

## Methods

### Sample collection and microscopy

Stool samples were collected within the framework of a multi-country randomized controlled trial assessing the efficacy and safety of the drug combination albendazole-ivermectin against *T. trichiura* and concomitant helminth infections [12]. The trial was approved by the Ethics Committee of Northwestern and Central Switzerland (EKNZ; BASEC Nr Req-2018-00494) and by the National Ethics Committee for Health Research in Lao PDR (No 093/NECHR). In the Nambak District where the study was conducted, Pak Khan village was selected for the daily sample collection for 28 days post-treatment based on previous good compliance and adequate number of participants living there. Prior to the start of collection, participants were informed of the aim of the daily sample collection, in addition to the consenting and information sessions conducted for the trial^33^. At the end of each sampling week, participants having provided at least one stool sample received a small gift of cooking oil. After collection, samples were transported to the field laboratory set up at the Nambak hospital, where they were analysed by standard Kato-Katz microscopy, which has been described elsewhere^33^. A small aliquot of < 1g stool was transferred to a 2ml screwcap cryotube using a UV-sterilised plastic spatula and immediately frozen at -20°C. After conclusion of the respective trial stage, the frozen samples were shipped directly to Swiss TPH on dry ice and kept at-20°C until analysis.

### DNA isolation

Samples were extracted using QIAamp DNA Mini kit (Qiagen, Hilden, Germany). The protocol used a garnet bead-beating approach in reference to a standard protocol developed by Kaisar *et al*^34^ with minor modifications. It has been described in more detail elsewhere^35^.

### 16S rRNA gene and shotgun sequencing

The V3-V4 hypervariable region of the 16S rRNA gene was sequenced as described previously^36^. Briefly, amplifications reactions were performed using 12.5 µl of Kapa HiFi HotStart ReadyMix (Roche, Basel, Switzerland) mix, 1.5 µl of 10 µM forward and reverse primers, 7.5 µl of sterile water, and 2 µl of template DNA. The V4 region was amplified by cycling the reaction at 95°C for 3 minutes, 30x cycles of 95°C for 15 seconds, 50°C for 15 seconds and 72°C for 15 seconds, followed by a 5 minutes 72°C extension. To avoid bias associated with PCR amplification, all reactions were done in triplicate, visually controlled on a 1.2% agarose and subsequently pooled together. Each pool was quantified using a Qubit hsDNA assay (Thermo Fisher Scientific, Waltham, MA, USA). The purified library was loaded onto an Illumina Miseq sequencer (Illumina, San Diego, CA, USA) as recommended by the manufacturer. Sequencing was performed using V3 chemistry (600 cycles) in 2x 300bp mode. Shallow shotgun sequencing was conducted as described^37^. Briefly, we generated a 4.4-13.3 M reads per sample to ensure representative taxonomic profiling with a better taxonomic resolution than what is allowed with 16S rRNA gene sequencing. DNA concentration was measured from 2 ul isolated DNA using a Qubit 4.0 in combination with hsDNA quantification kits (Thermo Fisher Scientific, Waltham, MA, USA). Libraries were prepared using the NEBNext Ultra II FS DNA kits (New England Biolabs, Ipswich, MA, USA) according to the manufacturers protocol. Final libraries were quantified and pooled together in an equimolar mix. The pool was subsequently loaded onto an Illumina NextSeq sequencer and sequenced using a Mid-output kit (300 cycles) in paired-end mode (2x 150bp). Sample sequencing depth is available in **Supplementary table 1**.

### Taxonomic profiling

The QIIME 2 pipeline v. 2020.8^38^ was used to analyse data generated by 16S rRNA gene sequencing. Briefly, after data import, Deblur was used to correct reads and cluster them into amplicon sequence variants with the option “--p-trim-length 140”. Taxonomic classification was done using the 16S rRNA gene database SILVA v.138^39^. Mitochondria and chloroplast related ASVs were removed using the “taxa filter-table” command. Alpha rarefaction curves were generated using “diversity alpha-rarefaction” command with “—p-max-depth 25000” option-Phylogenetic distances were calculated using the core-metrics-phylogenetic script and samples below 3000 classified reads were excluded using the option “–p-sampling-depth 3000”. Cleaned tables were exported and converted using the BIOM tool suite v. 2.1.9^40^. Metaphlan v. 3.0.7^41^ was used to perform taxonomic profiling on sequence data generated using the shallow shotgun sequencing technique. The species-markers database was CHOCOPhlAn v30 dating from January 2019 and reference mapping was done using Bowtie2 v. 2.4.5^42^. The option “—add_viruses” was added to the Metaphlan command to allow taxonomic profiling of viruses. Taxonomic profiles generated with Metaphlan3 were normalized by 16S qPCR values. A low count filter (minimum count = 100; prevalence = 20% of samples) and a low variance filter based on the inter-quantile range (50% features removed) were applied to the count table. Finally, data was transformed using the centered log ratio method^43^ before statistical analysis.

### qPCR analyses

qPCR assays were conducted on a CFX Opus real-time PCR system (Bio-Rad, Cressier, Switzerland) using Taqman Gene Expression analyses kits (Thermo Fisher Scientific, Waltham, MA, USA). qPCR targets included total bacteria (targeting the 16S gene)^44^, *Prevotella* genus^45^, *Faecalibacterium prausnitzii*^46^, and *Escherichia coli*^47^. Briefly, DNA was first diluted 10X in ultrapure water and each 10 µL reaction contained 0.3 µM of forward primer, 0.3 µM of reverse primer, and 0.2 µM (0.1 µM for *E. coli*) of Taqman probe, 5 µL of master mix, 2.2 µL of ultrapure water, and 2 µL of DNA template. Cycling conditions were as follows: 10 min at 95°C followed by 40 cycles at 95°C for 15 s and 60°C for 1 min. Detection threshold was set at 1500 RFU for targets detected with FAM, 700 RFU for HEX, and 200 RFU for CY5 and each reaction was conducted in duplicate. Primers and probes are summarised in **Supplementary table 2**.

### Statistical analysis

Group comparison using Mann-Whitney’s test, Fisher’s exact test of independence, and Spearman correlation tests were performed using XLSTAT v2020.2.3 (Addinsoft, New York, USA). Beta diversity measures and NMDS ordination plot derived from 16S rRNA gene sequencing were calculated on R v3.6.3^48^ with the following packages: vegan v2.5-6^49^ (with the vegdist, metaMDS) and hclust^50^. The package randomForest v4.6-14^51^ was used to run random forest models on 16S sequencing and qPCR data. The area under the receiving operator characteristic (AUROC) was generated using the pROC package v.1.16.2^52^. To identify enriched features in each enterotype, we used the LefSe package v.1.0^53^ (available on https://huttenhower.sph.harvard.edu/galaxy/). The survival analysis and cox proportional hazard were generated and measured using the “survfit” and “coxph”^54^ functions from the survival package, respectively. The heat tree analysis was performed on the MicrobiomeAnalyst platform^55^ using Metacoder R package^56^. All graphs, besides the heat tree, were generated using the OriginPro 2021 graphing software v9.8.0.200 (OriginLab Corporation, Northampton, MA, USA).

## Data Availability

16S rRNA gene and shotgun sequence data supporting the findings of this study have been deposited in the NCBI Short Read Archive with the primary accession code PRJNA767599.

## Author contributions

**PHHS**: study design, research design, project supervision, experimental work, statistical analyses, figure generation, writing of the initial manuscript, manuscript editing; **MG**: experimental work (16S rRNA gene sequencing, manuscript editing; **SW, EH, and SS**: study design, conducted field work (sample collection, handling, coordination of treatment, parasitological work, and data curation), experimental work (**SW**; DNA isolation), manuscript editing; **JD and CH**: experimental work (sample handling, DNA isolation, qPCR), manuscript editing; **JE**: experimental work (supervision of sequencing), manuscript editing; **JK**: study design, research design, project supervision, writing of the initial manuscript, manuscript editing.

## Acknowledgements

We thank Christian Beisel and Ina Nissen from the Genomics Facility Basel for their support during the shotgun sequencing experiments. Calculations were performed at sciCORE (https://scicore.unibas.ch) scientific computing center at the University of Basel on 40 cores and 480GB RAM.

